# Worldwide routine immunisation coverage regressed during the first year of the COVID-19 pandemic

**DOI:** 10.1101/2021.12.03.21267195

**Authors:** Beth Evans, Thibaut Jombart

## Abstract

We modelled historical, country-specific routine immunisation trends using publicly available vaccination coverage data for diphtheria, tetanus and pertussis-containing vaccine first-dose (DTP1) and third-dose (DTP3) from 2000 to 2019. We evaluate changes in coverage in 2020 by comparing model predictions to WUENIC-reported coverage. We report a 2.9% (95%_CI_: [2.2%; 3.6%]) global decline in DTP3 coverage, and important increases in missed immunisations in some countries with middle-income countries, and the Americas, being most affected.

---

The COVID-19 pandemic has impacted society and public health infrastructures worldwide, influencing mobility [1], access to health services [2], livelihoods and poverty [3]. While COVID-19 vaccination strategies continue to receive considerable emphasis [4,5], the extent to which routine immunisation (RI) has been impacted during the first year of the pandemic remains unclear. Indeed, the World Health Organisation (WHO) pulse surveys reported disruptions in the first half of 2020 [2], and while some later studies suggested a potential recovery [6], recent observations again hinted at global coverage declines [7].

RI is estimated to prevent four to five million deaths worldwide every year [8]. As such, there is an urgent need for assessing potential changes in RI coverage, as declines may result in considerable added morbidity and mortality.

## A global decline in RI coverage

We investigated changes in RI coverage using two key indicators: diphtheria-tetanus-pertussis first-dose (DTP1) and third-dose (DTP3). DTP3 serves as a general marker for immunisation system performance, used by national and global immunisation stakeholders [9]. DTP1 is used as a proxy for inequity - quantifying Zero Dose (ZD) children, those that receive no childhood vaccinations [10].

We compiled vaccination coverage data from the WHO and United Nations Children’s Fund (UNICEF) Estimates of National Immunisation Coverage (WUENIC) [11,12] for the last 20 years. We used AutoRegressive Integrated Moving Average (ARIMA) modelling [13] to capture temporal trends in coverage for each country from 2000 to 2019, and predicted expected coverage levels in 2020 (**Figure 1**). All analyses were conducted using R [14] and can be reproduced using a publicly available *reportfactory [15]* including all required data and scripts [16].

**Figure 1:**
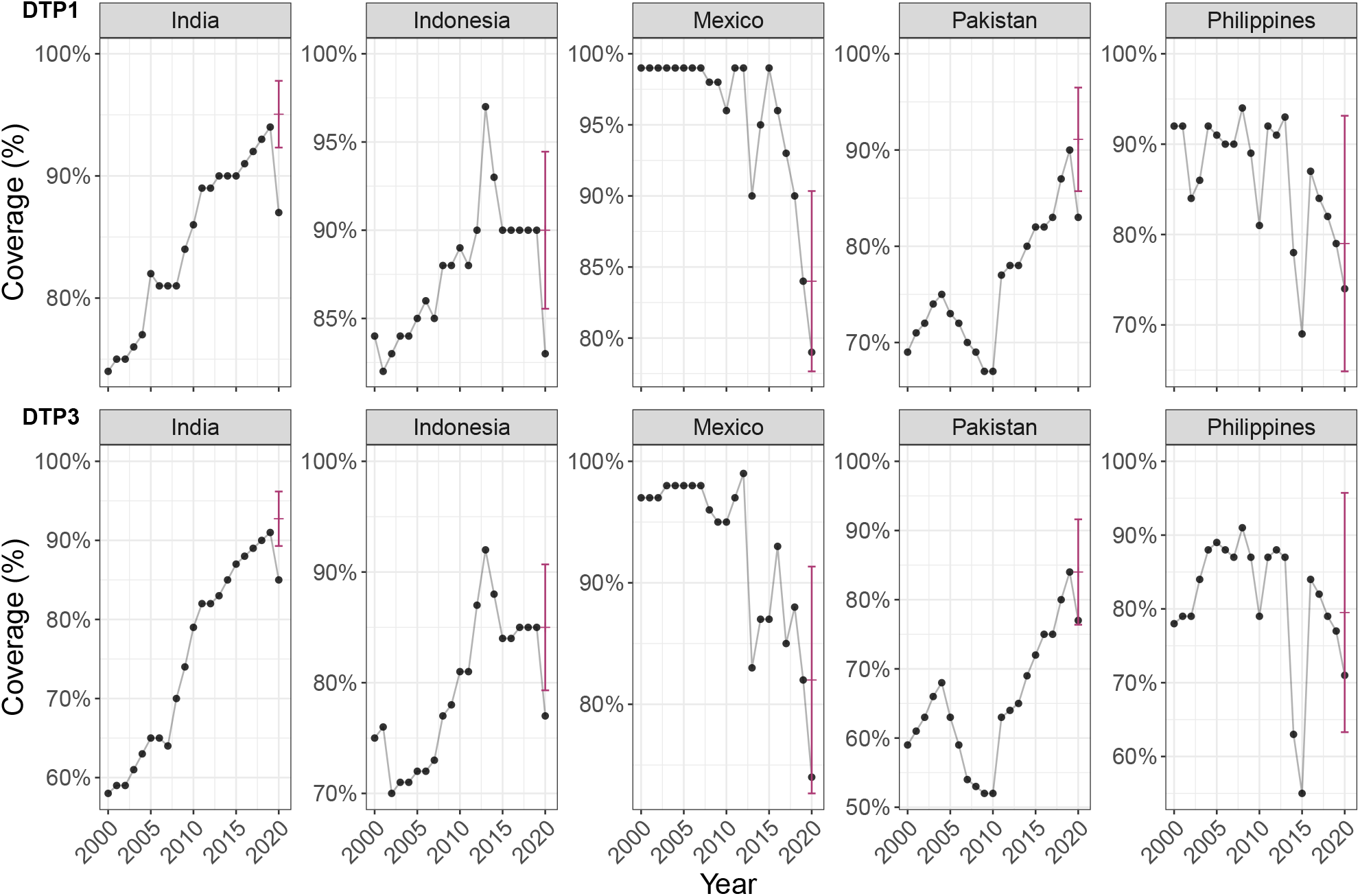
Expected and reported 2020 vaccine coverage for DTP1 and DTP3: example of five countries with most additional missed DTP3 immunisations in 2020. These graphs show WUENIC-reported coverage data (black dots) and the corresponding ARIMA predictions and the associated 95% confidence intervals (red bars).

After excluding countries for which reliable coverage predictions could not be obtained (see Supplementary Text for details), we were able to estimate differences between expected and observed coverage in 2020 for 167 countries for DTP3 (**Figure 2**) and DTP1. Unfortunately, the exact magnitude of coverage decline was often hard to assess for individual countries due to uncertainties in model predictions (**Figure 2, Supplementary Table 1 and 2**), but general trends could still be identified.

**Figure 2:**
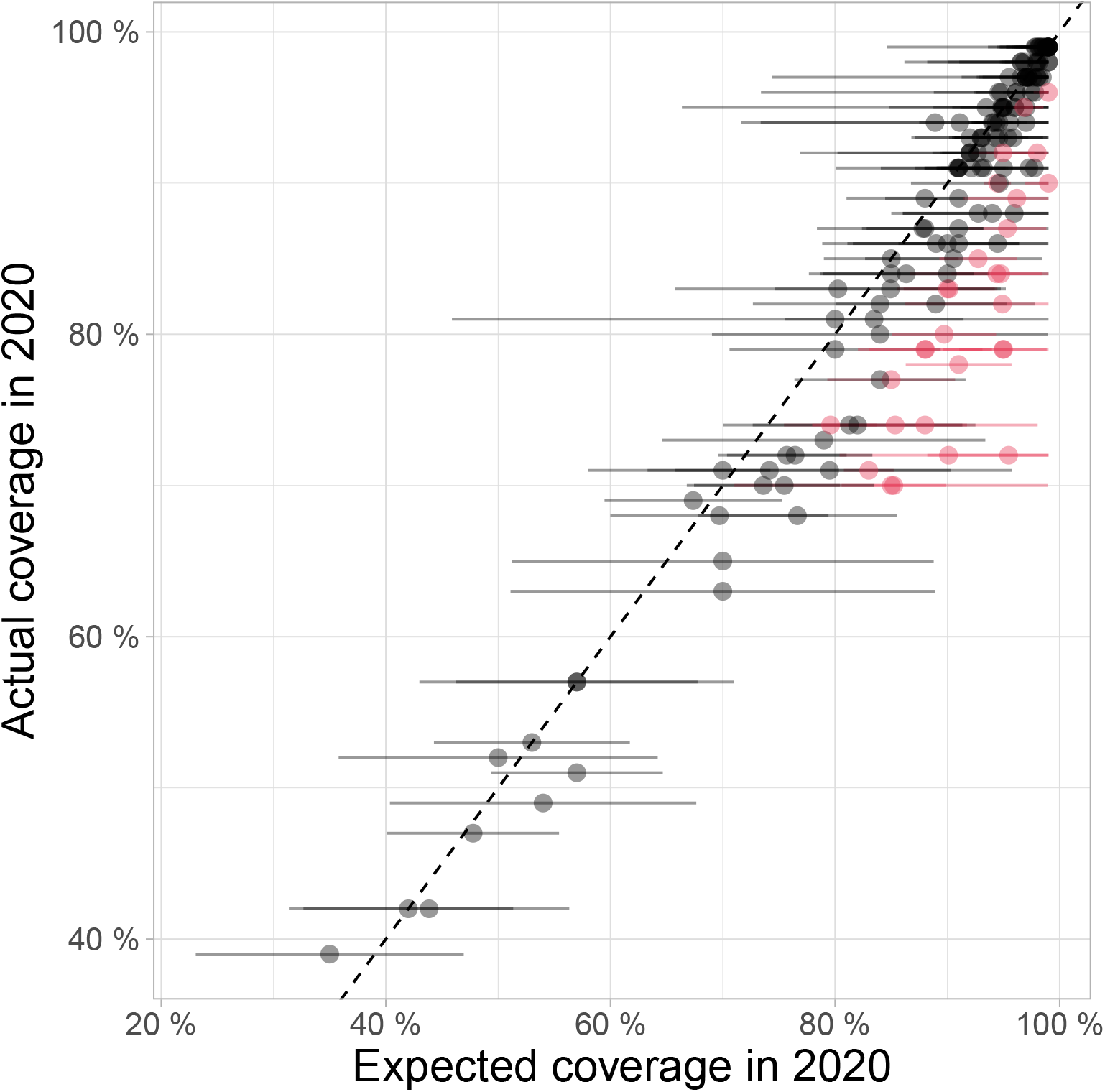
Comparison between 2020 WUENIC-reported DTP3 coverage and expectations derived from historical trends. This scatterplot shows country coverage (WUENIC-reported actuals and ARIMA-predicted expectations) as dots. Lines around individual points illustrate the 95% confidence intervals (CI) of ARIMA predictions. Countries showing significant departure from expected values, *i*.*e*., for which actual coverage is outside the 95% CI of predictions, are indicated in red.

Results suggest an average global decline in DTP3 coverage of 2.9% (95%_CI_: [2.2%; 3.6%]), from an expected 89.2% to a reported 86.3% across 167 reporting countries. While this change may seem small, such levels of coverage was last observed in these countries in 2005, thus suggesting a potential 15-years setback in RI improvements.

Similar trends were seen for DTP1 - an average global coverage decline of 2.2% (95%_CI_: [1.6%; 2.8%]) from an expected 92.9% to a reported 90.7% across the 167 countries analysed here. Declines in DTP1 coverage indicate increases in the quantity of ZD children in some countries - suggesting that the most vulnerable populations have been strongly impacted by the reductions in RI observed in the first year of the pandemic and reinforced existing inequities in access to healthcare.

## Heterogeneities in RI disruption

While the global decline in DTP3 coverage is in itself an important result, it is also essential to characterise potential differences among affected countries. Whilst individual country estimates have uncertainty, larger-scale investigations of heterogeneities could be achieved by comparing groups of countries (**Figure 3**). Indeed, patterns of RI coverage significantly varied across United Nations regions (**Figure 3A**; ANOVA: *F* = 22.4, df = 162, *p* < 2.2×10^−16^), with the strongest decline observed in the Americas (6.2% decline, 95%_CI_: [4.6%; 7.7%]), Asia (3.5% decline, 95%_CI_: [2.2%; 4.8%]) and Africa (2.8% decline, 95%_CI_: [1.6%; 4.0%]), while Europe (mean change = -0.6%; 95%_CI_: [-2.0%; +0.7%]) and Oceania (mean change = -0.4%; 95%_CI_: [-2.9%; +2.2%]) did not show any significant change.

**Figure 3:**
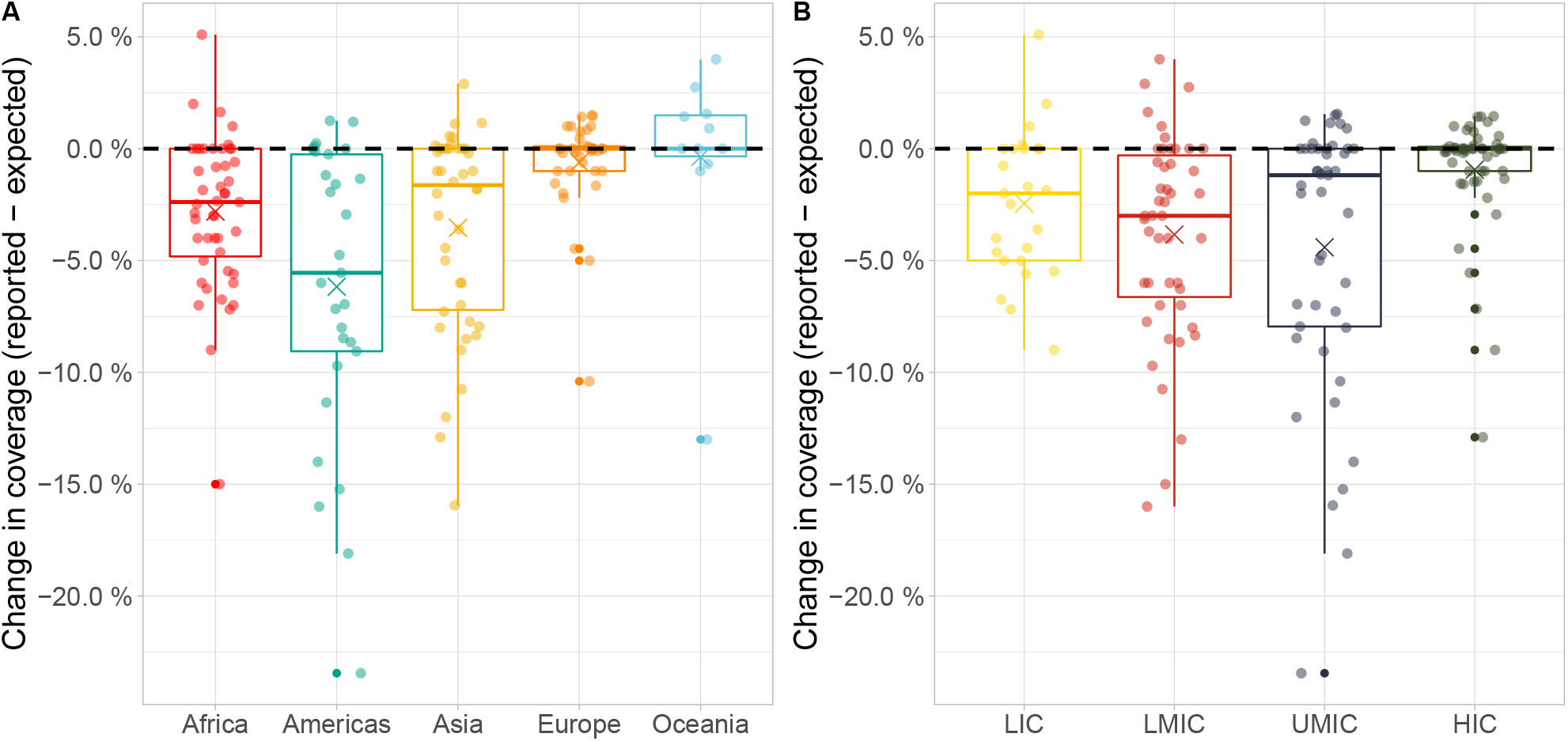
Differences between expected and reported DTP3 vaccine coverage in 2020 across (A) UN regions and (B) income groups. Points represent individual countries, grouped, and coloured according to (A) UN region classification and (B) World Bank income groups. Country coordinates on the X-axis were jittered for visibility. Values on the y-axis are indicated as absolute differences between reported and expected vaccine coverage, in percentages. Boxes show the median (50%), upper (75%) and lower (25%) quartile changes in coverage for each group, with whiskers extending to either the minimum/ maximum changes or the quartile value plus 1.5 times the interquartile range, and crosses indicating the average. The black dashed horizontal lines indicate no change in coverage. LIC: Low-income Country. LMIC: Lower-middle-income Country. UMIC: Upper-middle-income Country. HIC: High-income Country

Similar heterogeneities were observed when considering income groups (**Figure 3B**; ANOVA: F = 22.6, df = 163, p < 7.1^-15^), with stronger declines in coverage observed in lower middle income countries (LMICs; mean decline: 3.8%; 95%_CI_: [2.6%; 5.1%]) and in upper middle income countries (UMICs; mean decline: 4.4% ; 95%_CI_: [3.1%; 5.7%]), than in low income countries (LICs; mean decline: 2.4%; 95%_CI_: [0.7%; 4.2%]), while high income countries (HICs; mean change: -0.9%; 95%_CI_: [-2.2%; 0.3%]) did not show any significant change.

As UN regions and income groups are highly correlated (non-parametric Chi-square test: *X*^2^= 115.4, p < 10^−5^), we also tested whether heterogeneities due to one variable (regions or income groups) remained after accounting for the effect of the other one. Interestingly, regional differences remained after accounting for differences in income groups (ANOVA: *F* = 5.67, df = 159, p < 2.7×10^−4^), but evidence for the converse was weak (ANOVA: *F* = 2.67, df = 159, p = 0.05).

Combined with surviving infant estimates (medium variant births minus infant deaths) of the United Nations World Population Prospects (UNWPP) for 2020 [17], our results suggest a strong impact, with large additional missed immunisations versus expected, in some countries. For example, in India there were an estimated 3.5 M unvaccinated children for DTP3 in 2020, of which 52% (95%CI: [29%; 75%]) were associated with the pandemic disruption; and in Indonesia an estimated 1.1 M missed DTP3 vaccinations, of which 35% (95%_CI_: [10%; 60%]) were associated with coverage declines in 2020. **Table 1** details results for the 10 countries with point estimate greatest additional missed DTP3 immunisations in 2020 (see also **Figure 1**). Similar trends are seen for ZD children using DTP1 results.

**Table 1:**
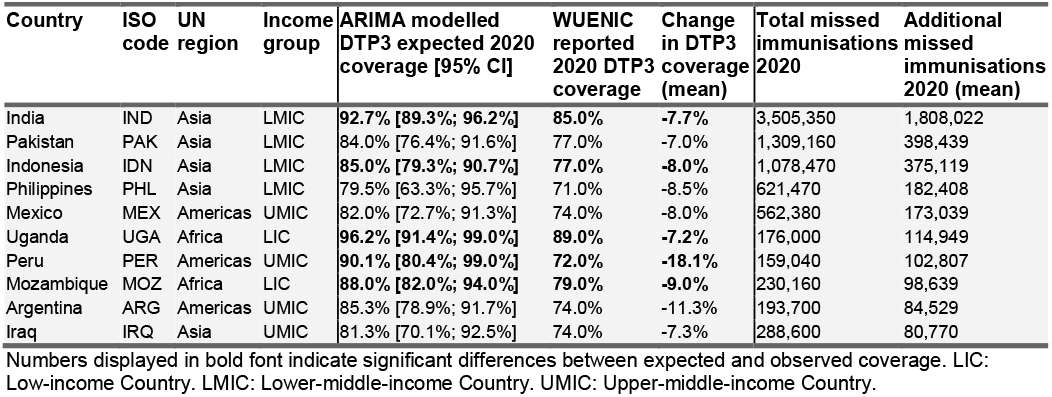
Estimated DTP3 coverage declines and missed immunisations for 10 countries with most additional missed immunisations.

| Country | ISO code | UN region | Income group | ARIMA modelled DTP3 expected 2020 coverage [95% CI] | WUENIC reported 2020 DTP3 coverage | Change in DTP3 coverage (mean) | Total missed immunisations 2020 | Additional missed immunisations 2020 (mean) |
| --- | --- | --- | --- | --- | --- | --- | --- | --- |
| India | IND | Asia | LMIC | **92.7% [89.3%; 96.2%]** | **85.0%** | **-7.7%** | 3,505,350 | 1,808,022 |
| Pakistan | PAK | Asia | LMIC | 84.0% [76.4%; 91.6%] | 77.0% | -7.0% | 1,309,160 | 398,439 |
| Indonesia | IDN | Asia | LMIC | **85.0% [79.3%; 90.7%]** | **77.0%** | **-8.0%** | 1,078,470 | 375,119 |
| Philippines | PHL | Asia | LMIC | 79.5% [63.3%; 95.7%] | 71.0% | -8.5% | 621,470 | 182,408 |
| Mexico | MEX | Americas | UMIC | 82.0% [72.7%; 91.3%] | 74.0% | -8.0% | 562,380 | 173,039 |
| Uganda | UGA | Africa | LIC | **96.2% [91.4%; 99.0%]** | **89.0%** | **-7.2%** | 176,000 | 114,949 |
| Peru | PER | Americas | UMIC | **90.1% [80.4%; 99.0%]** | **72.0%** | **-18.1%** | 159,040 | 102,807 |
| Mozambique | MOZ | Africa | LIC | **88.0% [82.0%; 94.0%]** | **79.0%** | **-9.0%** | 230,160 | 98,639 |
| Argentina | ARG | Americas | UMIC | 85.3% [78.9%; 91.7%] | 74.0% | -11.3% | 193,700 | 84,529 |
| Iraq | IRQ | Asia | UMIC | 81.3% [70.1%; 92.5%] | 74.0% | -7.3% | 288,600 | 80,770 |

Numbers displayed in bold font indicate significant differences between expected and observed coverage. LIC: Low-income Country. LMIC: Lower-middle-income Country. UMIC: Upper-middle-income Country.

Detailed results for all analysed countries can be found in the **Supplementary Tables S1** (DTP1) and **S2** (DTP3).

## Discussion

The estimated changes in RI coverage reported in this study suggest a smaller global decline (approximately 1/3rd the magnitude) than previously found using alternative methodology and data [18]. We believe our findings may be more robust owing to a more comprehensive dataset including data from more countries (167 here vs. 94), plus increased data from the end of 2020 (annual here vs. majority of data from January-September 2020), and the use of WUENIC-reported data (less prone to data quality and completeness issues than administrative data). The observed discrepancies are compatible with a rebound of global RI coverage in late 2020 [6].

The RI disruption observed in this study suggests there may be greater risk of vaccine-preventable disease outbreaks in the coming years, in the absence of Supplementary Immunisation Activities (SIAs) to reach missed children. ZD populations in key ZD “hotspots” (*e*.*g*., India, Pakistan, and Indonesia) are estimated to have increased significantly in 2020, posing a genuine public health threat in the coming years. To alleviate such risks and reduce immunisation inequities, SIAs targeted specifically at these populations should be considered. Additional research is needed to investigate heterogeneities in RI decline at finer scales and identify subpopulations which may have experienced even greater losses to RI coverage.

RI disruption may be worsened by the acceleration of COVID-19 vaccination campaigns, particularly in low- and middle-income countries where absorption capacity may be challenged [19,20], potentially competing with RI services. Careful monitoring of the interaction, trade-offs and synergies between RI and COVID-19 vaccinations is essential. Further studies are needed to understand which factors linked to the COVID-19 crisis impacted vaccination coverage, such as changes in health-seeking behaviours or non-pharmaceutical intervention policies, in order to successfully and efficiently address pandemic-associated losses to coverage.

As the COVID-19 pandemic continues to affect healthcare systems globally, maintaining the appropriate balance between access to routine immunization and pandemic response will be essential to reduce both the direct and indirect mortality and morbidity associated with COVID-19. This research provides a transparent and replicable rationale for estimating gaps in RI coverage across countries, producing an objective measure for missed immunisations and coverage disruptions. As such, it can form a basis for identifying countries most affected by declines in RI coverage and prioritising efforts to alleviate the indirect impact of COVID-19.

## Data Availability

All data produced in the present study use publicly available datasets. All data and reproducible analyses are available online at-
https://github.com/bevans249/modelling_covid_imact_RI

https://doi.org/10.5281/zenodo.5750111

## Notes

### Competing Interest Statement

The authors have declared no competing interest.

### Funding Statement

TJ receives funding from the Global Challenges Research Fund (GCRF) project RECAP managed through RCUK and ESRC (ES/P010873/1), from the National Institute for Health Research - Health Protection Research Unit for Modelling Methodology, and from the Medical Research Council (grant number MC_PC_19065). These funders had no role in the design and conduct of this study; collection, management, analysis, and interpretation of the data; preparation, review, or approval of the manuscript; and decision to submit the manuscript for publication.
BE is an MSc student at the London School of Hygiene and Tropical Medicine, and received no funding.

