## Supplementary Text for "Worldwide routine immunisation coverage regressed during the first year of the COVID-19 pandemic"

**Additional data, methodological information and complementary results**

*This supplementary material is hosted by Eurosurveillance as supporting information alongside the article “Worldwide routine immunisation coverage regressed during the first year of the COVID-19 pandemic” on behalf of the authors who remain responsible for the accuracy and appropriateness of the content. The same standards for ethics, copyright, attributions and permissions as for the article apply. Eurosurveillance is not responsible for the maintenance of any links or email addresses provided therein."*

**Contents:**

Section S1: Vaccination coverage data

Section S2: Further datasets for categorising countries

Section S3: Using ARIMA to model expected vaccination coverage

Section S4: Countries removed from analyses

Section S5: Full results

Section S6: Availability and reproducibility

**Section S1: Vaccination coverage data**

Vaccine coverage is typically estimated through (a) aggregating (raw) administration data, or (b) conducting surveys. Expert opinion and/ or statistical methods may be layered on to produce final estimates. Administration data is the most timely and periodic, but risks numerator (e.g., under- or over-estimation dependent on health system capacity, reporting incentives, and linkage to private sector systems) and denominator (e.g., out-of-date censuses used for population quantification) biases [[1]](https://paperpile.com/c/NLQygk/MHhw). Surveys, typically household or parental, avoid such biases but are expensive, time-consuming, infrequent, and may encounter recall bias, i.e., parents or guardians may mis-remember or confuse vaccination statuses when reporting due to complex immunisation schedules and potentially long periods between delivery and surveys [[2]](https://paperpile.com/c/NLQygk/IJ2x). The Institute for Health Metrics and Evaluation (IHME) produces coverage estimates by applying spatiotemporal statistical methods to household survey microdata (if available) and estimates of country-reported coverage data [[3]](https://paperpile.com/c/NLQygk/pzb5). IHME usefully publish confidence ranges, unlike other sources, but methods are not fully reproducible nor used routinely by global immunisation stakeholders to assess immunisation performance. WUENIC estimates are published annually through computational logic rule-based approaches that use inputs from country-reported administrative and survey data, adjusted based on expert assessment and country consultations [[4,5]](https://paperpile.com/c/NLQygk/doI1+pxVm). WUENIC estimates are transparent, replicable, and routinely used by stakeholders to inform policy, financial, and programmatic decisions.

WUENIC data was selected due to its transparency, replicability, utility by key organisations and donors working on or investing in immunisation globally (e.g., UNICEF, WHO and Gavi), and public availability. From this data, two RIs – DTP1 and DTP3 – were selected since they act as key immunisation indicators. DTP1 is typically delivered 6-weeks after birth [[6]](https://paperpile.com/c/NLQygk/gFqk), and is used as a proxy for inequity, specifically quantifying the number of Zero Dose children [[7]](https://paperpile.com/c/NLQygk/733g). DTP3 is typically administered between 14-weeks and 22-weeks after birth [[6]](https://paperpile.com/c/NLQygk/gFqk), and is a key indicator of immunisation programmer performance. It can also be used as a proxy for fully immunised child coverage – children that received all recommended basic vaccines by 12-months of age [[8]](https://paperpile.com/c/NLQygk/mK2a).

**Section S2: Further datasets for categorising countries**

Three additional sources were used to assemble country demographic information – population data from the United Nations World Population Prospects (UNWPP, [33]), World Bank (WB) income group classification [34], and United Nations (UN) regional classifications (using the “countrycode” package in R). The final list of sources – all publicly available online – are summarised in **Supplementary Table 2**.

**Section S3: Using ARIMA to model expected vaccination coverage**

WUENIC does not publish future-looking coverage forecasts. Expected coverage for 2020, by country in the absence of COVID-19 was modelled by fitting AutoRegressive Integrated Moving Average (ARIMA) models to the last 20 years of annual WUENIC coverage data (2000-2019) using the package “*forecast*”) in R [[9]](https://paperpile.com/c/NLQygk/WkDL) and projecting models forward one year. These predicted 2020 coverage estimates are referred to as “expected coverage” from here-on. No projections were modelled for countries missing any coverage values from 2000 to 2019.

For each time series the appropriate ARIMA model based on historic coverage trends was automatically selected using the function “*auto.arima*” in the “*forecast”* package [[9]](https://paperpile.com/c/NLQygk/WkDL). ARIMA models are characterised by three order terms – *p*, *d*, and *q* – representing the order of the Auto Regressive (AR) term, the number of differencing steps required to make the time series stationary (Integrated, I, term), and the order of the Moving Average (MA) term respectively. Since WUENIC data has annual periodicity, there are no seasonal patterns, and seasonal ARIMA models were not required. Automatic ARIMA modelling fits the appropriate *p*, *d*, and *q* order terms based on:

●	Conducting Kwiatkowski-Phillips-Schmidt-Shin tests, with the null hypothesis that the time series is stationary around a deterministic trend against the alternative of a unit root to validate whether the time series is stationary [[10]](https://paperpile.com/c/NLQygk/42Uj). Where not stationary, each time series is differenced and then re-validated to test if stationary to determine the integrative order, *d*.

●	Stepwise algorithm to traverse the model space to select the model with the smallest Akaike Information Criterion (AIC) [[11]](https://paperpile.com/c/NLQygk/rZ7s). AIC scores evaluate how well a model fits the data it was generated from, and lower AIC scores (for the same dataset) indicate better model fit. AIC minimisation identifies the *p* and *q* terms.

The mean ARIMA-modelled value was selected per country and vaccine as expected 2020 coverage. WUENIC cap coverage at 99% [[4,5]](https://paperpile.com/c/NLQygk/doI1+pxVm). To avoid falsely calculating small declines in coverage for countries where ARIMA-predicted 2020 coverage was higher than 99%, all expected coverage estimates were capped at the WUENIC maximum.

**Section S4: Countries removed from analyses**

Before further analysis, coverage time series including expected coverage were investigated to identify unreliable estimates based on meeting one or more of the following criteria:

1)	**Volatility**: WUENIC coverage estimates showing fluctuations greater than 10 percentage points (pps) over the last decade, indicating high unreliability and uncertainty in point estimates

2)	**Strong influence of recent potentially anomalous estimates**: Visual inspection of fitted models revealed high dependency on major coverage increases/decreases in a single year (2018 or 2019). Such data points may have over contributed to model fitting, but reflect exceptional circumstances (e.g., stockouts, civil war)

3)	**High modelled coverage improvement:** Predicted improvements of greater than or equal to five pps between reported 2019 coverage and expected 2020 coverage, which may not be programmatically feasible, unless 2019 was justified as an exception with coverage expected to return to “normal”

Where countries met any of these criteria, country-specific WUENIC coverage documentation was reviewed to assess data quality and relevant context [[12]](https://paperpile.com/c/NLQygk/ko71). Countries were removed from further analysis where WUENIC documentation suggested data was highly uncertain (e.g., administration data does not match survey data, or many recent revisions), or recent data reflected exceptional circumstances, such as temporary stockouts, or political unrest.

The following countries were removed for the following reasons for DTP1 analyses:

- **Criteria one - Volatility**
  - Solomon Islands: Coverage exhibits high variance over time. WUENIC reported that the "increase in estimated coverage from 2018 [to 2019] is unlikely but exceptionally allowed given the small birth cohort size" [[12]](https://paperpile.com/c/NLQygk/ko71).
  - Suriname: Recent coverage trends appear volatile. WUENIC reported that "recent survey results suggest lower levels of coverage than that reported by the programme during the past 10 years. Further investigation to understand underlying differences is warranted, and WHO and UNICEF recommend a high-quality independent empirical assessment to confirm reported levels of coverage" [[12]](https://paperpile.com/c/NLQygk/ko71). WUENIC also reported stockouts in 2019 and 2017 (one month), syringe stockout in 2018 (four months), which may have impacted achievable coverage.
- **Criteria two - Strong influence of recent potentially anomalous estimates:**
  - Haiti: Model fit skewed heavily by 2019 coverage estimate, which fell due to socio-political disturbances according to WUENIC documentation [[12]](https://paperpile.com/c/NLQygk/ko71).
  - Libya: Model fit skewed heavily by 2018-2019 coverage estimates, which were all reduced 25 pps vs. country reports based on reported 3-month stockout, and therefore are not strongly tied to surveys.
  - Samoa: Model fit skewed heavily by 2018-2019 coverage estimates (much lower than historical trend). WUENIC noted vaccine interruption in 2018 due public hesitancy, and that 2019 data is uncertain.
  - Brazil: Forecast driven by fit to 2019 coverage, which was greater than 15 pps reduction in a single year. WUENIC reported "no nationally representative household survey within the last 5 years" so unclear whether anomalous or indicative of future trends [[12]](https://paperpile.com/c/NLQygk/ko71).
- **Criteria three - High forecast coverage improvement:**
  - Jordan: +7 pp improvement on 2019 coverage may be programmatically unlikely. Furthermore between 2018 and 2019 there was a 10 pp reported decrease in coverage (not reported as a one-off exception), adding increased volatility and uncertainty to recent estimates.
  - Bolivia: +6 pp improvement forecast. WUENIC coverage estimates reported recent uncertainty due to civil unrest [[12]](https://paperpile.com/c/NLQygk/ko71).
  - Austria: +5 pp improvement forecast. Country did not report data in 2019 so the 2019 WUENIC estimate was based on extrapolation [[12]](https://paperpile.com/c/NLQygk/ko71).

The following countries were removed for the following reasons for DTP3 analyses:

- **Criteria one - volatility**:
  - Solomon Islands: Coverage exhibits high variance over time. WUENIC reported that "increase in estimated coverage from 2018 [to 2019] is unlikely but exceptionally allowed given the small birth cohort size" [[12]](https://paperpile.com/c/NLQygk/ko71).
  - Suriname: Coverage exhibits recent volatility. WUENIC report "recent survey results suggest lower levels of coverage than that reported by the programme during the past 10 years. Further investigation to understand underlying differences is warranted, and WHO and UNICEF recommend a high-quality independent empirical assessment to confirm reported levels of coverage" as well as stock outs in 2019 and 2017 (one month), syringe stockout in 2018 (four months) [[12]](https://paperpile.com/c/NLQygk/ko71).
- **Criteria two - Strong influence of recent potentially anomalous estimates:**
  - Haiti: Model fit skewed heavily by 2019 coverage estimate, which fell due to socio-political disturbances according to WUENIC documentation [[12]](https://paperpile.com/c/NLQygk/ko71).
  - Libya: Model fit skewed heavily by 2018-2019 coverage estimates. These estimates were reduced by a flat 25 pps during the WUENIC review process due to country reports based on reported 3-month stockout (however survey results remain higher) [[12]](https://paperpile.com/c/NLQygk/ko71).
  - Samoa: Model fit skewed heavily by 2018-2019 coverage estimates (much lower than historical trends). WUENIC noted vaccine interruption in 2018 due public hesitancy, and that 2019 data is uncertain [[12]](https://paperpile.com/c/NLQygk/ko71). Samoa also meets criteria three – with point estimate improvement of +6 pp from 2019 levels predicted.
  - Brazil: Model estimate driven by fit to 2019 coverage which was > 15 pp drop in a year), and WUENIC report "no nationally representative household survey within last 5 years" so unclear how trend will evolve or if it will be confirmed [[12]](https://paperpile.com/c/NLQygk/ko71).
- **Criteria three - High forecast coverage improvement:**
  - Venezuela: +6 pp improvement may be programmatically unlikely. WUENIC coverage estimates reported uncertainty due to civil unrest [[12]](https://paperpile.com/c/NLQygk/ko71).
  - El Salvador: +6 pp improvement may be programmatically unlikely. WUENIC reported no survey in the last 5 years and a need for high-quality survey [[12]](https://paperpile.com/c/NLQygk/ko71).
  - Jordan: +7 pp improvement on 2019 coverage may be programmatically unlikely. Furthermore between 2018 and 2019 there was a 10 pp reported decrease in coverage (not reported as a one-off exception [[12]](https://paperpile.com/c/NLQygk/ko71)), adding increased volatility and uncertainty to recent estimates.

**Section S5: Full results**

Full output results are available in **Supplementary Materials Table S1** for DTP1, and **Supplementary Materials Table S2** for DTP3. These tables include:

- Country classification details: country, ISO code, UN region and income group
- ARIMA predictions: mean, low- and high- 95% confidence intervals, confidence interval width
- WUENIC reported 2020 coverage: from WUENIC
- Deltas (expected - reported coverage): mean, low- and high- 95% confidence intervals
- 95% confident flag: binary flag whether WUENIC-reported 2020 coverage is outside the ARIMA-predicted 95% confidence intervals
- Surviving infant population: from UN WPP medium variant births minus infant mortality
- Missed immunisation estimates: total missed immunisations (based on WUENIC-reported 2020 coverage), Expected (based on mean ARIMA prediction coverage)
- Additional missed immunisation estimates: mean (based on mean coverage delta), low and high estimates (based on delta 95% confidence intervals)

**Section S6: Availability and reproducibility**

All analyses were conducted using R 4.1.1 [[13]](https://paperpile.com/c/NLQygk/zV7N). All the data and R code implementing the analyses are publicly available from the following github repository: <https://github.com/bevans249/modelling_covid_imact_RI>
